# A Conversational Artificial Intelligence Framework for Comparative Pathway-Level Profiling of Sézary Syndrome and Primary Cutaneous CD8^+^ Aggressive Epidermotropic Cytotoxic T-Cell Lymphoma (PCAECTCL)

**DOI:** 10.64898/2026.04.15.26350992

**Authors:** Fernando C. Diaz, Brigette Waldrup, Francisco G. Carranza, Sophia Manjarrez, Enrique Velazquez-Villarreal

## Abstract

**Background:** Sézary syndrome (SS) is an aggressive leukemic variant of cutaneous T-cell lymphoma (CTCL) with distinct clinical and biological features compared to rarer entities such as primary cutaneous CD8⁺ aggressive epidermotropic cytotoxic T-cell lymphoma (PCAECTCL). Although recurrent genomic alterations in CTCL have been described, comparative analyses at the pathway level across biologically divergent subtypes remain limited. Here, we leveraged a conversational artificial intelligence (AI) platform for precision oncology to enable rapid, integrative, and hypothesis-driven interrogation of publicly available genomic datasets.

**Methods:** We conducted a secondary analysis of somatic mutation and clinical data from the Columbia University CTCL cohort accessed via cBioPortal. Cases were stratified into SS (n=26) and PCAECTCL (n=13). High-confidence coding variants were curated and mapped to biologically relevant signaling pathways and functional gene categories implicated in CTCL pathogenesis. Pathway-level mutation frequencies were compared using Chi-square or Fisher’s exact tests, with effect sizes quantified as odds ratios. Tumor mutational burden (TMB) was compared using the Wilcoxon rank-sum test. Subtype-specific co-mutation patterns were evaluated using pairwise association analyses and visualized through oncoplots and network heatmaps. Conversational AI agents, AI-HOPE, were used to iteratively refine cohort definitions, prioritize pathway-level signals, and contextualize findings.

**Results:** TMB was comparable between SS and PCAECTCL (p = 0.96), indicating no significant difference in global mutational load. In contrast, pathway-centric analyses revealed marked qualitative differences. SS demonstrated enrichment of alterations in epigenetic regulators, tumor suppressor and cell-cycle control pathways, NFAT signaling, and DNA damage response mechanisms, consistent with transcriptional dysregulation and immune modulation. PCAECTCL exhibited relatively higher frequencies of alterations involving epigenetic regulators and MAPK pathway signaling, suggesting distinct oncogenic dependencies. Co-mutation analysis revealed a more constrained and focused interaction landscape in SS, whereas PCAECTCL displayed broader and more heterogeneous co-mutation networks, indicative of divergent evolutionary trajectories. Notably, ERBB2 mutations were significantly enriched between subtypes (p = 0.031), highlighting a potential subtype-specific therapeutic vulnerability.

**Conclusions:** This study demonstrates that SS is distinguished from PCAECTCL not by increased mutational burden but by distinct pathway-level architectures, particularly involving epigenetic regulation, immune signaling, and transcriptional control. These findings generate biologically grounded, testable hypotheses for subtype-specific therapeutic targeting and underscore the value of conversational AI as a scalable framework for accelerating discovery in translational cancer genomics.

## 1. Introduction

Cutaneous T-cell lymphoma (CTCL) comprises a biologically diverse group of extranodal non-Hodgkin lymphomas characterized by the clonal expansion of malignant skin-homing T lymphocytes. Although rare, CTCL represents a clinically significant malignancy due to its chronic, often relapsing course, diagnostic complexity, and potential for progression to systemic disease. Population-based analyses estimate that more than 14,000 individuals were diagnosed with CTCL in the United States between 2000 and 2018, with incidence trends gradually increasing over time (1). Despite accounting for only ∼4% of non-Hodgkin lymphomas and ∼0.14% of all cancers (2–6), CTCL poses a disproportionate clinical burden, underscoring the need for improved molecular characterization and more effective, biology-driven therapeutic strategies (7–9).

CTCL encompasses multiple clinicopathologic entities with distinct clinical behaviors and outcomes. Among these, Sézary syndrome (SS) represents a prototypical aggressive leukemic variant, defined by erythroderma, lymphadenopathy, and circulating malignant T cells, and is associated with poor prognosis (7). In contrast, primary cutaneous CD8⁺ aggressive epidermotropic cytotoxic T-cell lymphoma (PCAECTCL) is a rare but highly aggressive cytotoxic CTCL subtype characterized by rapid progression, epidermotropism, and often fulminant clinical course. While both entities exhibit aggressive phenotypes, their clinical presentation, cellular origin, and therapeutic responses differ substantially, suggesting fundamentally distinct biological underpinnings. Importantly, CTCL subtypes frequently mimic benign inflammatory dermatoses, including eczema and psoriasis, particularly in early stages, contributing to delayed diagnosis and suboptimal clinical management (8,10).

The management of CTCL remains challenging and requires a multidisciplinary approach integrating dermatologic, hematologic, and oncologic expertise. Therapeutic strategies are highly heterogeneous and stage-dependent, ranging from skin-directed therapies to systemic immunomodulatory agents, targeted therapies, and radiation (7,8,10,11). Although recent advances, including immune checkpoint inhibitors and monoclonal antibodies such as mogamulizumab, have expanded the therapeutic landscape, durable responses remain limited for many patients (12,13). This therapeutic variability highlights a critical need for improved molecular stratification to guide precision oncology approaches tailored to specific CTCL subtypes.

At the molecular level, CTCL pathogenesis is complex and incompletely understood, involving the interplay of somatic genomic alterations, dysregulated signaling pathways, and tumor microenvironment interactions. Prior genomic studies have identified recurrent mutations affecting key oncogenic pathways, including T-cell receptor signaling, MAPK and RAS signaling, epigenetic regulation, and cytokine-mediated pathways such as JAK-STAT (14–19). Integrated genomic analyses further demonstrate substantial molecular heterogeneity across CTCL subtypes, supporting the concept that distinct genetic and signaling programs drive divergent clinical phenotypes (16,20). More recent single-cell and spatial transcriptomic studies have revealed complex tumor ecosystems, including TH2-skewed malignant T-cell populations interacting with immunosuppressive microenvironments enriched in B cells and regulatory immune circuits (21–23). Together, these findings emphasize that CTCL biology is governed not only by individual gene alterations but by coordinated dysregulation of signaling networks within a dynamic microenvironment.

Despite these advances, most genomic investigations in CTCL have focused on individual gene mutations or subtype-restricted analyses, with limited efforts directed toward systematic pathway-level comparisons across biologically distinct entities such as SS and PCAECTCL. Given that oncogenesis is driven by network-level perturbations rather than isolated genetic events, pathway-centric frameworks may provide a more mechanistic and clinically actionable understanding of subtype-specific disease biology.

Recent developments in artificial intelligence (AI) have introduced new paradigms for accelerating discovery in translational cancer genomics. AI-driven analytical systems enable scalable interrogation of complex, multidimensional datasets, facilitating pattern recognition, hypothesis generation, and integrative interpretation beyond traditional analytical approaches (24,25). In particular, conversational AI platforms have emerged as flexible tools that allow dynamic, natural language-guided exploration of clinical and genomic data, enabling iterative refinement of analytical hypotheses and rapid identification of biologically relevant signals.

In this study, we leveraged AI-HOPE (26), a conversational AI framework designed for precision oncology, along with pathway-specific modules including AI-HOPE-JAK-STAT (27) and AI-HOPE-MAPK (28), to perform a pathway-centric comparative analysis of CTCL subtypes. AI-HOPE enables dynamic cohort construction, pathway-level aggregation of genomic alterations, and interactive exploration of molecular relationships within large-scale datasets such as cBioPortal. By integrating AI-guided analysis with conventional statistical methods, this framework supports both robust quantitative evaluation and biologically informed interpretation of complex genomic landscapes (29,30).

Using this approach, we conducted a secondary analysis of the Columbia University CTCL cohort to systematically compare the somatic mutational architecture of SS and PCAECTCL (3,31–35). Through pathway-level profiling and gene-gene interaction analyses, we aimed to identify subtype-specific molecular signatures and signaling dependencies that distinguish these aggressive CTCL entities. By integrating pathway-centric genomics with conversational AI-enabled exploration, this work seeks to advance understanding of CTCL heterogeneity and provide a foundation for the development of more precise, subtype-informed therapeutic strategies.

## 2. Methods

### 2.1 Data Source and Cohort Definition

This study represents a secondary analysis designed to molecularly characterize and compare the somatic mutational landscape of SS and PCAECTCL using publicly available genomic and clinical datasets. Mutation and clinical annotation data were obtained from the Columbia University CTCL cohort accessible through the cBioPortal for Cancer Genomics platform. cBioPortal provides harmonized genomic datasets derived from multiple institutional studies and allows standardized analysis of somatic mutation and clinical features across cancer cohorts.

Tumor samples were stratified into two groups based on the detailed cancer type annotation within the dataset. The SS consisted of samples annotated specifically as SS (n = 26). The PCAECTCL cohort consisted of sample size of 13. All samples were classified within the broader disease category of mature T- and NK-cell neoplasms. This stratification enabled direct comparison of genomic alterations between the leukemic CTCL subtype SS and PCAECTCL.

### 2.2 Mutation Data Processing and Filtering

Somatic mutation data were retrieved directly from the cBioPortal mutation annotation files associated with the Columbia CTCL dataset. To focus analyses on biologically meaningful alterations with potential functional consequences, only high-impact coding variants were retained for downstream analyses. High-impact variants were defined according to variant classification categories commonly associated with functional protein changes. These included: Missense mutations, Nonsense mutations, Frameshift insertions and deletions, Splice-site mutations, In-frame insertions and deletions, and Translation start-site alterations

This filtering strategy enriched the dataset for variants most likely to influence protein structure, gene function, and downstream signaling pathways relevant to CTCL biology.

### 2.3 Gene-Level and Pathway-Level Mutation Annotation

Filtered somatic variants were annotated to predefined functional gene groups and signaling pathways based on established biological roles in CTCL pathogenesis and T-cell malignancies. Gene group assignments were curated from previously reported CTCL genomic studies (3) and canonical pathway annotations derived from established biological databases (3).

The following functional gene groups and signaling pathways were analyzed: epigenetic regulators (TET2, DNMT3A, CREBBP, KMT2D, KMT2C, ARID1A, SMARCA4, CHD3, and BRD9), which are involved in chromatin remodeling, DNA methylation, and transcriptional regulation; tumor suppressor genes (TP53, RB1, PTEN, CDKN1B, and CDKN2A), critical for maintaining genomic stability, regulating cell-cycle checkpoints, and controlling apoptotic signaling; cell-cycle regulators (TP53, RB1, CDKN1B, and CDKN2A), which govern key checkpoints in cell-cycle progression; T-cell receptor (TCR) signaling components (PLCG1, VAV1, LAT, LCK, ZAP70, and FYN), mediating proximal TCR activation and downstream signaling cascades; the JAK-STAT signaling pathway (JAK1, JAK3, STAT3, STAT5B, and SOCS1), which regulates cytokine-mediated signaling and immune activation; the MAPK signaling pathway (MAPK1, MAPK3, BRAF, KRAS, NRAS, RAF1, MAP2K1, MAP2K2, NF1, and RASA1), representing central nodes of the mitogen-activated protein kinase cascade; the NF-κB signaling pathway (CARD11, TNFAIP3, IKBKB, CHUK, NFKB1, NFKB2, REL, RELA, and RELB), which controls inflammatory signaling and T-cell survival; the NFAT signaling pathway (NFATC1, NFATC2, PRKG1, PPP3CA, PPP3CB, and PPP3R1), involved in calcium-dependent transcriptional activation and immune signaling; DNA damage response genes associated with genomic integrity and DNA repair processes, including TP53 and related components; and genes involved in apoptosis and immune regulation (FAS, FASLG, BCL2, BCL6, PDCD1, and CTLA4), which regulate programmed cell death, immune checkpoint signaling, and tumor immune evasion.

For each tumor sample, a functional gene group or signaling pathway was classified as mutated if at least one high-impact somatic variant was present in any gene belonging to that pathway. Pathway mutation status was therefore represented as a binary variable (mutated vs. wild type) for downstream statistical comparisons between cohorts.

### 2.4 Pathway-Level Statistical Analysis

Mutation frequencies for each functional gene group and signaling pathway were compared between the SS and PCAECTCL cohorts. Statistical comparisons were performed using Chi-square or Fisher’s exact tests, which is appropriate for categorical comparisons in small sample sizes.

To quantify the magnitude and direction of associations between pathway alterations and CTCL subtype, odds ratios (ORs) and corresponding confidence intervals were estimated. Results were visualized using forest plots to facilitate interpretation of pathway-level enrichment patterns across CTCL subtypes.

Gene-level mutation frequencies were also calculated for each cohort, and Chi-square or Fisher’s exact tests was used to identify genes with potential subtype-specific enrichment. Genes demonstrating borderline statistical significance were highlighted for exploratory interpretation and hypothesis generation.

### 2.5 Tumor Mutation Burden Analysis

Tumor mutation burden (TMB) values were obtained directly from the clinical annotation files provided in the cBioPortal dataset. TMB represents the total number of somatic mutations per tumor sample and serves as a surrogate measure of global mutational load.

TMB distributions between SS and PCAECTCL cohorts were compared using boxplot visualization and statistical testing with the Wilcoxon rank-sum test. This nonparametric test was selected because it does not assume normal distribution of mutation counts and is appropriate for small cohort sizes.

### 2.6 Co-Mutation Analysis

To evaluate subtype-specific patterns of mutational interaction, pairwise gene-gene co-mutation analyses were performed using the most frequently mutated genes in the dataset. Analyses were conducted separately for the SS and PCAECTCL cohorts to identify patterns of mutational co-occurrence or mutual exclusivity.

Statistical testing of gene-gene associations was performed using Chi-square or Fisher’s exact tests. Results were visualized as co-mutation heatmaps, enabling identification of recurrent interaction patterns among mutated genes within each CTCL subtype.

### 2.7 Visualization of Mutational Landscape

Multiple visualization approaches were used to summarize the genomic landscape of CTCL subtypes: Oncoplots were generated to display somatic mutation patterns across the most frequently altered genes in the cohort. Each column represents an individual tumor sample and each row represents a gene. Colored blocks indicate the presence and classification of high-impact mutations. Lollipop plots were generated for the top recurrently mutated genes in each cohort to illustrate mutation positions along the protein sequence and identify potential mutation hotspots. Forest plots were used to summarize pathway-level mutation enrichment and associated odds ratios between cohorts.

### 2.8 Conversational Artificial Intelligence-Assisted Analysis

A conversational AI platform developed for precision oncology research - AI-HOPE (26), AI-HOPE-JAK-STAT (27) and AI-HOPE-MAPK (28) - was used to facilitate interactive exploration, prioritization, and interpretation of genomic findings. The AI framework enabled rapid querying of genomic datasets, dynamic visualization of mutation patterns, and generation of pathway-level hypotheses by integrating mutation data, biological pathway annotations, and statistical outputs.

This AI-assisted workflow served as a complementary analytical layer to conventional statistical methods, accelerating hypothesis generation and enabling efficient interpretation of complex genomic data while maintaining transparency and reproducibility of analytical steps.

## 3. Results

### 3.1 Cohort composition and subtype-specific classification

The analytical cohort consisted of 39 CTCL tumors derived from the Columbia University dataset available through cBioPortal, stratified into two biologically and clinically distinct groups: Sézary syndrome (SS; n = 26) and primary cutaneous CD8⁺ aggressive epidermotropic cytotoxic T-cell lymphoma (PCAECTCL; n = 13) (Table 1). All cases were classified within the overarching category of mature T- and NK-cell neoplasms.

**Table 1.**
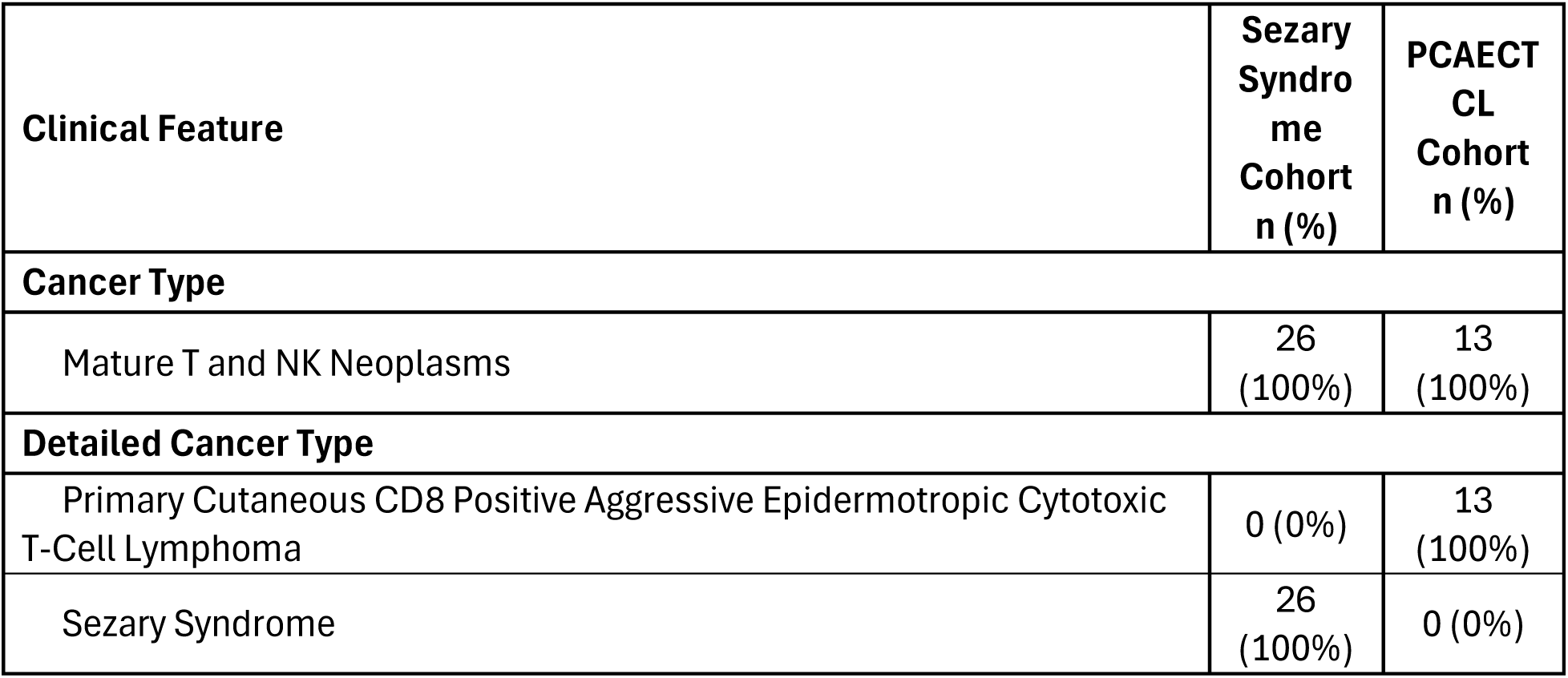
Clinical and histopathologic characteristics of the Sézary syndrome and PCAECTCL cohorts.

| Clinical Feature | Sézary Syndrome Cohort n (%) | PCAECTCL Cohort n (%) |
| --- | --- | --- |
| **Cancer Type** |  |  |
| Mature T and NK Neoplasms | 26 (100%) | 13 (100%) |
| **Detailed Cancer Type** |  |  |
| Primary Cutaneous CD8 Positive Aggressive Epidermotropic Cytotoxic T-Cell Lymphoma | 0 (0%) | 13 (100%) |
| Sézary Syndrome | 26 (100%) | 0 (0%) |

This study employed a focused binary stratification design to enable direct comparison between two aggressive CTCL entities with distinct immunophenotypic and clinical characteristics. The SS cohort predominantly represents leukemic CD4⁺ T-cell-driven disease, whereas the PCAECTCL cohort is characterized by CD8⁺ cytotoxic epidermotropic lymphomas, a biologically and clinically aggressive subtype.

This subtype-restricted framework reduces biological heterogeneity and enhances interpretability of downstream pathway-level analyses, enabling clearer delineation of molecular differences associated with T-cell lineage (CD4⁺ versus CD8⁺), disease behavior, and underlying oncogenic signaling programs. By focusing on these two aggressive CTCL variants, the cohort design provides a robust foundation for identifying subtype-specific molecular architectures and signaling dependencies.

### 3.2 Comparable tumor mutational burden in Sézary syndrome and PCAECTCL cohorts

We first examined whether SS was distinguished from PCAECTCL by differences in overall mutational burden (Fig. 1). TMB obtained from the cBioPortal clinical annotations, showed substantial inter-sample variability in both groups, but the overall distributions were highly comparable. Median TMB values and interquartile ranges overlapped extensively between SS and PCAECTCL, and Wilcoxon rank-sum testing demonstrated no significant difference between cohorts (P = 0.96). These data indicate that the molecular distinction between SS and PCAECTCL is not driven by a higher global mutational load in SS, but instead is more likely attributable to qualitative differences in the identity and pathway context of somatic alterations.

**Figure 1.**
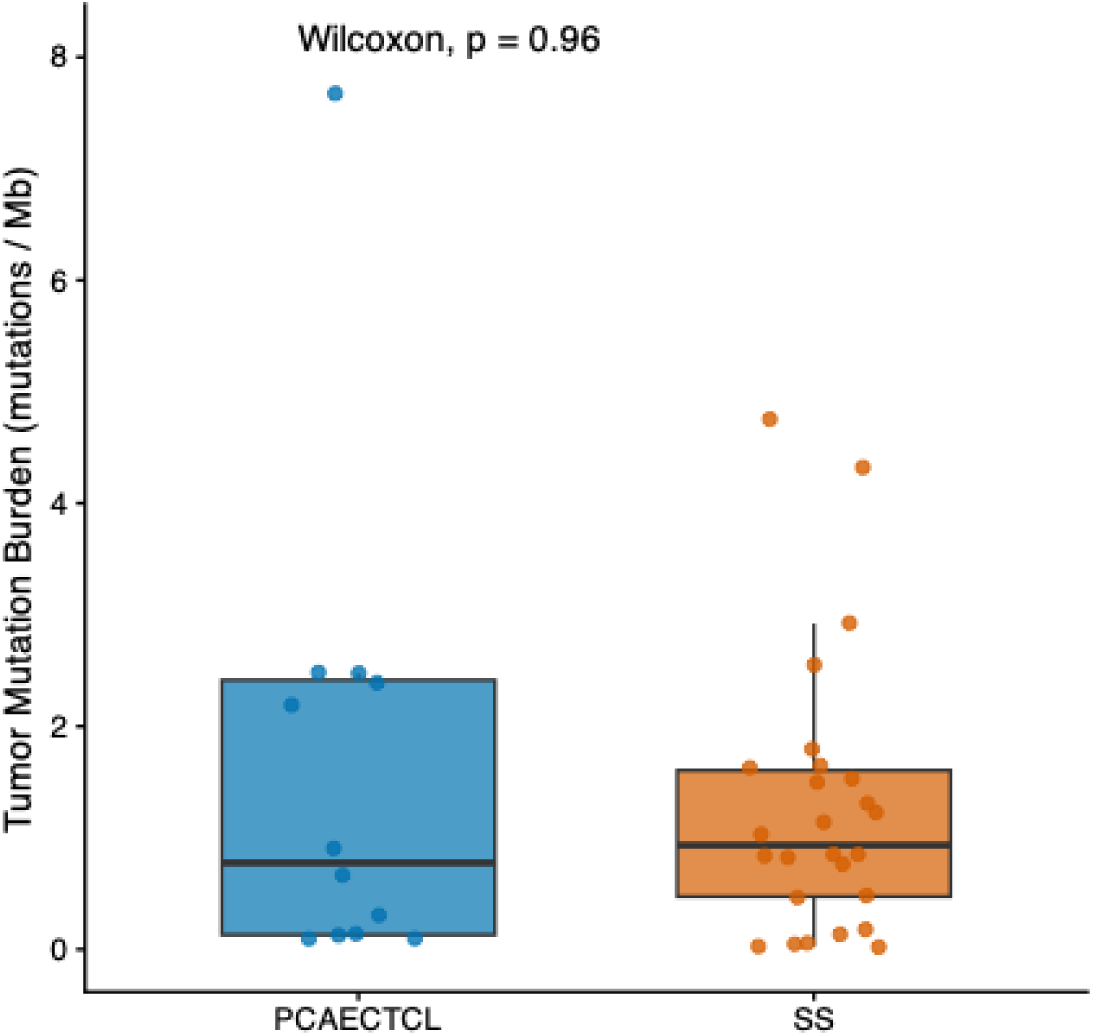
Comparable tumor mutational burden between Sézary syndrome and PCAECTCL.

### 3.3 Divergent pathway architectures distinguish Sézary syndrome from PCAECTCL

To define subtype-specific biological programs, we mapped high-impact somatic variants to curated signaling pathways and functional gene categories relevant to T-cell lymphomagenesis (Table 2). Comparative pathway-level analysis revealed a consistent divergence between SS and PCAECTCL, highlighting distinct patterns of pathway engagement rather than differences in overall mutational burden.

**Table 2.** Pathway-level comparison of somatic mutation frequencies between Sézary syndrome and PCAECTCL.

| Functional Group or Pathway | Sézary Syndrome Mutated n (%) | Sézary Syndrome Wild-Type n (%) | PCAECTCL Mutated n (%) | PCAECTCL Wild-Type n (%) | p-value |
| --- | --- | --- | --- | --- | --- |
| Epigenetic Regulation | 10 (38%) | 16 (62%) | 3 (23%) | 10 (77%) | 0.4774 |
| Tumor Suppressor Pathway | 4 (15%) | 22 (85%) | 1 (8%) | 12 (92%) | 0.6478 |
| Cell Cycle Regulation | 4 (15%) | 22 (85%) | 1 (8%) | 12 (92%) | 0.6478 |
| T-Cell Receptor Signaling | 3 (12%) | 23 (88%) | 0 (0%) | 13 (100%) | 1 |
| JAK/STAT Signaling | 1 (4%) | 25 (96%) | 1 (8%) | 12 (92%) | 1 |
| MAPK Signaling | 2 (8%) | 24 (92%) | 3 (23%) | 10 (77%) | 0.3102 |
| NF-κB Signaling | 2 (8%) | 24 (92%) | 1 (8%) | 12 (92%) | 1 |
| NFAT Signaling | 4 (15%) | 22 (85%) | 1 (8%) | 12 (92%) | 0.6478 |
| DNA Damage Response | 4 (15%) | 22 (85%) | 1 (8%) | 12 (92%) | 1 |
| Apoptosis and Immune Regulation | 1 (4%) | 25 (96%) | 0 (0%) | 13 (100%) | 1 |

SS demonstrated a broader involvement of pathways related to transcriptional regulation and immune signaling. Alterations affecting epigenetic regulators were identified in 38% of SS tumors compared with 23% of PCAECTCL cases, representing the most prominent pathway-level enrichment in SS. Similarly, tumor suppressor genes and cell-cycle regulatory pathways were altered in 15% of SS tumors versus 8% in PCAECTCL, suggesting a greater contribution of canonical growth control disruption in the SS phenotype. Pathways associated with T-cell receptor (TCR) signaling were exclusively altered in SS (12% vs. 0%), further supporting the notion of aberrant antigen receptor-driven signaling as a defining feature of this leukemic subtype. In addition, NFAT signaling alterations were more frequent in SS (15% vs. 8%), consistent with dysregulated transcriptional programs linked to T-cell activation and differentiation.

Alterations in DNA damage response pathways and apoptosis/immune regulatory mechanisms also showed a trend toward enrichment in SS, with the latter observed only in SS tumors (4% vs. 0%), albeit at low frequency. In contrast, NF-κB signaling alterations were detected at comparable rates across both subtypes (8%), suggesting a shared role in CTCL biology rather than subtype-specific dependency.

PCAECTCL, by contrast, exhibited relatively higher involvement of MAPK signaling, with mutations observed in 23% of cases compared to 8% in SS. JAK/STAT pathway alterations were present at low and comparable frequencies in both subtypes (4% in SS vs. 8% in PCAECTCL), indicating that this axis may not represent a primary distinguishing feature between these two aggressive CTCL entities.

Although none of the pathway-level comparisons reached statistical significance, likely reflecting the limited cohort size, the directionality and magnitude of differences across multiple pathways suggest biologically meaningful divergence. Specifically, SS appears to be characterized by preferential perturbation of epigenetic, transcriptional, and immune-regulatory circuits, whereas PCAECTCL demonstrates a relative shift toward MAPK-driven oncogenic signaling.

Together, the observed patterns indicate that Sézary syndrome and PCAECTCL are governed by fundamentally different pathway-level organizations, likely shaped by their distinct cellular origins, immune microenvironments, and underlying oncogenic drivers (Figure S1). Framing these differences at the level of signaling networks, rather than individual genes, offers a more comprehensive lens to interpret CTCL heterogeneity and may inform the development of more precise, subtype-adapted therapeutic strategies.

### 3.4 Gene-level comparison reveals subtype-specific mutational signatures and differential driver distribution

To refine the molecular distinctions observed at the pathway level, we performed a detailed gene-level comparison of somatic alterations between Sézary syndrome (SS) and PCAECTCL cohorts (Table 3). This analysis highlighted contrasting mutational architectures, with SS exhibiting a more recurrent and subtype-restricted pattern, while PCAECTCL demonstrated a broader but less concentrated distribution of genetic alterations.

**Table 3.** Gene-level comparison of somatic mutation frequencies between Sézary syndrome and PCAECTCL cohorts.

| Gene | Sézary Syndrome Mutated n (%) | Sézary Syndrome Wild-Type n (%) | PCAECTCL Mutated n (%) | PCAECTCL Wild-Type n (%) | p-value |
| --- | --- | --- | --- | --- | --- |
| STAT3 | 0 (0%) | 26 (100%) | 1 (8%) | 12 (92%) | 0.3333 |
| NFKB1 | 0 (0%) | 26 (100%) | 1 (8%) | 12 (92%) | 0.3333 |
| TET2 | 3 (12%) | 23 (88%) | 0 (0%) | 13 (100%) | 0.5377 |
| NFATC2 | 0 (0%) | 26 (100%) | 1 (8%) | 12 (92%) | 0.3333 |
| KMT2D | 0 (0%) | 26 (100%) | 1 (8%) | 12 (92%) | 0.3333 |
| MAPK1 | 0 (0%) | 26 (100%) | 2 (15%) | 11 (85%) | 0.1053 |
| LCK | 0 (0%) | 26 (100%) | 0 (0%) | 13 (100%) | 1 |
| JAK3 | 0 (0%) | 26 (100%) | 0 (0%) | 13 (100%) | 1 |
| SH2B3 | 0 (0%) | 26 (100%) | 0 (0%) | 13 (100%) | 1 |
| BRAF | 0 (0%) | 26 (100%) | 1 (8%) | 12 (92%) | 0.3333 |
| BRCA1 | 0 (0%) | 26 (100%) | 0 (0%) | 13 (100%) | 1 |
| BRCA2 | 0 (0%) | 26 (100%) | 0 (0%) | 13 (100%) | 1 |
| ATR | 0 (0%) | 26 (100%) | 2 (15%) | 11 (85%) | 0.3333 |
| PLCG1 | 3 (12%) | 23 (88%) | 0 (0%) | 13 (100%) | 0.5377 |
| ARID1A | 0 (0%) | 26 (100%) | 2 (15%) | 11 (85%) | 0.1053 |
| PREX2 | 0 (0%) | 26 (100%) | 2 (15%) | 11 (85%) | 0.1053 |
| TP53 | 3 (12%) | 23 (88%) | 0 (0%) | 13 (100%) | 0.5377 |
| CDKN2A | 0 (0%) | 26 (100%) | 1 (8%) | 12 (92%) | 0.3333 |
| PRKG1 | 2 (8%) | 24 (92%) | 0 (0%) | 13 (100%) | 0.5439 |
| SMARCA4 | 1 (4%) | 25 (96%) | 1 (8%) | 12 (92%) | 1 |
| STAT5B | 1 (4%) | 25 (96%) | 0 (0%) | 13 (100%) | 1 |
| CREBBP | 2 (8%) | 24 (92%) | 0 (0%) | 13 (100%) | 0.5439 |
| CHD3 | 2 (8%) | 24 (92%) | 0 (0%) | 13 (100%) | 0.5439 |
| BRD9 | 1 (4%) | 25 (96%) | 0 (0%) | 13 (100%) | 1 |
| CARD11 | 2 (8%) | 24 (92%) | 0 (0%) | 13 (100%) | 0.5439 |
| NF1 | 1 (4%) | 25 (96%) | 0 (0%) | 13 (100%) | 1 |
| MAPK3 | 1 (4%) | 25 (96%) | 0 (0%) | 13 (100%) | 1 |
| PDCD1 | 1 (4%) | 25 (96%) | 0 (0%) | 13 (100%) | 1 |
| KMT2C | 1 (4%) | 25 (96%) | 1 (8%) | 12 (92%) | 1 |

In the SS cohort, the most frequently mutated genes were TET2, PLCG1, and TP53, each identified in 12% of cases (3/26). These recurrent alterations were accompanied by additional events in PRKG1, CREBBP, CHD3, and CARD11 (each 8%), as well as lower-frequency mutations in STAT5B, BRD9, NF1, MAPK3, and PDCD1. This constellation of mutations reflects a convergence on epigenetic regulation, T-cell receptor signaling, transcriptional modulation, and immune checkpoint pathways, consistent with a tightly organized and biologically coherent mutational program underlying SS.

In contrast, PCAECTCL displayed a more heterogeneous mutational profile with fewer recurrent events and greater dispersion across genes. The most frequent alterations were observed in MAPK1, ARID1A, PREX2, and ATR (each 15%), suggesting a relative enrichment of MAPK pathway signaling, chromatin remodeling, and DNA damage response processes. Additional single-occurrence mutations were identified in genes such as STAT3, NFKB1, NFATC2, KMT2D, BRAF, and CDKN2A, indicating involvement of multiple signaling pathways without a clear dominant driver. Notably, several genes commonly implicated in signaling cascades, including LCK, JAK3, and SH2B3, were not mutated in either cohort, underscoring selective pathway engagement rather than ubiquitous activation.

A key observation was the presence of subtype-restricted mutations. Alterations in TET2, PLCG1, and TP53 were exclusive to SS, reinforcing their potential role as defining molecular features of this subtype. Conversely, mutations in MAPK1, ARID1A, PREX2, ATR, and several signaling mediators were observed only in PCAECTCL, suggesting distinct oncogenic dependencies. Only a limited number of genes, such as SMARCA4 and KMT2C, were shared between both groups at low frequencies, indicating minimal overlap in core mutational drivers.

Although statistical significance was not achieved for individual gene comparisons, likely due to cohort size constraints, the overall distribution patterns provide meaningful biological insight. SS appears to be driven by recurrent alterations in a defined set of genes linked to regulatory and immune-related functions, whereas PCAECTCL exhibits a more fragmented mutational landscape spanning multiple oncogenic pathways.

Altogether, these gene-level distinctions reinforce the concept that SS and PCAECTCL are molecularly divergent entities. Focusing on individual gene alterations not only refines subtype classification but also highlights candidate targets for future functional validation and precision therapeutic development.

We next evaluated genes demonstrating subtype-associated enrichment at the individual gene level (Table S1). ERBB2 emerged as a notable discriminatory candidate, with mutations detected in 3 of 13 PCAECTCL tumors (23%) and in none of the 26 SS tumors, yielding a statistically significant difference (p = 0.03129). This pattern suggests that ERBB2 alterations may represent a subtype-associated molecular feature of PCAECTCL rather than a shared CTCL event. Given the established role of ERBB2 as a receptor tyrosine kinase with well-described oncogenic and therapeutic relevance across multiple malignancies, its selective enrichment in PCAECTCL is of particular interest. Although this finding requires validation in larger independent cohorts, it raises the possibility that ERBB2-associated signaling may contribute to PCAECTCL biology and could represent a candidate vulnerability for future precision-targeted investigation.

### 3.5 Subtype-specific gene-gene interaction networks reveal divergent mutational organization

To further interrogate the structure of somatic alterations beyond individual genes and pathways, we evaluated pairwise gene-gene interaction patterns within each cohort, focusing on the top 20 most frequently mutated genes in Sézary syndrome (SS) and PCAECTCL (Fig. 2). This analysis uncovered clear differences in the organization and complexity of mutational interactions between the two subtypes.

**Figure 2.**
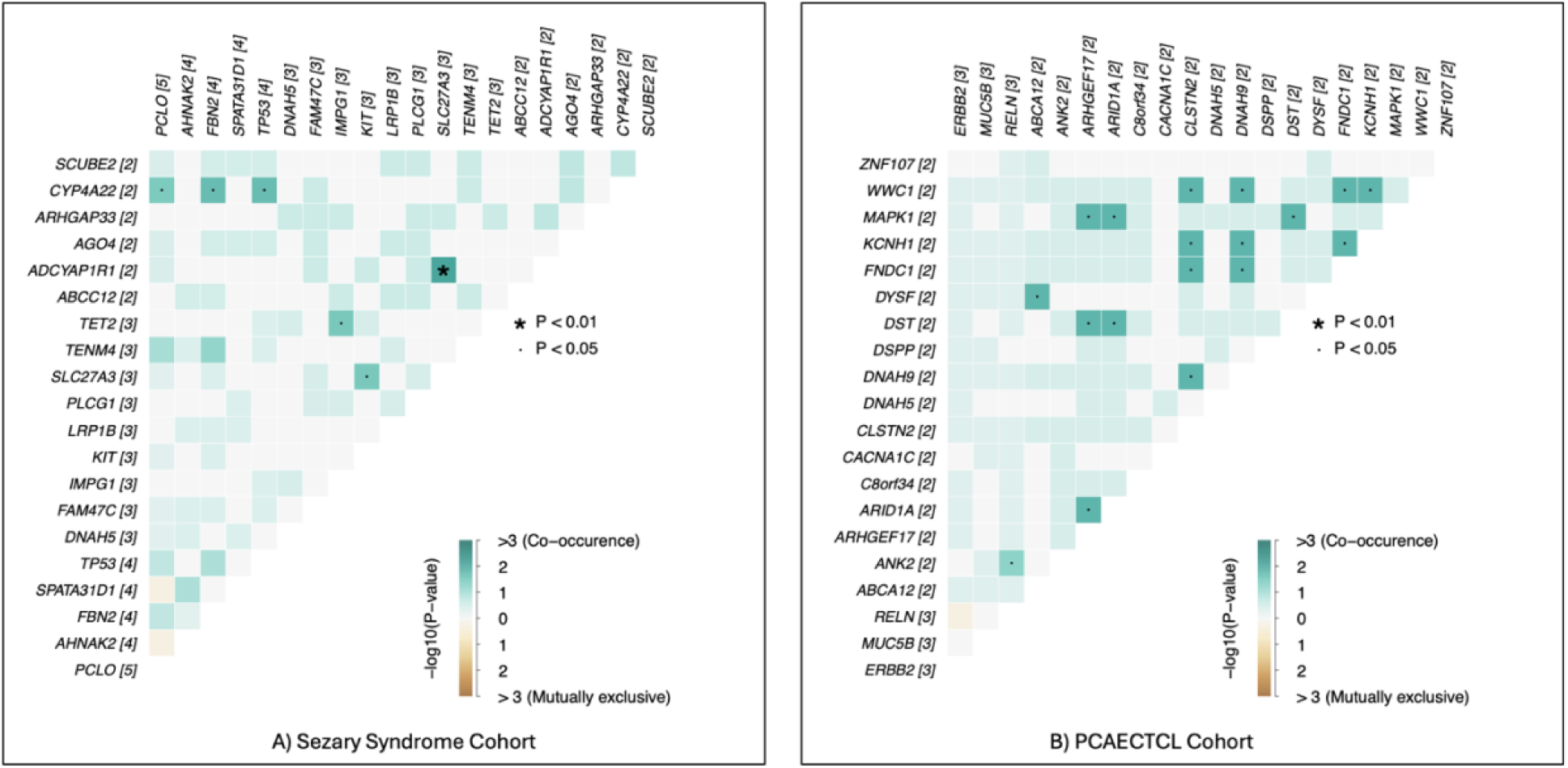
Distinct gene-gene mutational interaction patterns in Sézary syndrome and PCAECTCL cohorts. Subtype-stratified gene-gene co-mutation heatmaps were constructed for the 20 most frequently mutated genes in Sézary syndrome (SS) and Primary Cutaneous CD8⁺ Aggressive Epidermotropic Cytotoxic T-Cell Lymphoma (PCAECTCL) cohorts. Pairwise mutational interactions were evaluated to identify statistically significant co-occurrence or mutual exclusivity relationships across tumor samples. The SS cohort (panel A) demonstrated a relatively limited interaction network, with six significant gene-gene associations detected. In contrast, the PECACTCL cohort (panel B) exhibited a substantially larger set of significant interactions (18 gene pairs), indicating a broader mutational co-occurrence landscape. Heatmap color intensity reflects the −log10(p-value) of pairwise associations, with positive values representing co-occurrence and negative values indicating mutual exclusivity; asterisks denote statistically significant interactions (P < 0.01), and dots indicate nominal significance (P < 0.05). These results highlight subtype-specific mutational architectures in CTCL, with PCAECTCL tumors displaying more complex gene-gene interaction patterns compared with the relatively constrained mutational network observed in SS.

In the SS cohort, the co-mutation landscape was relatively sparse, with a limited number of statistically supported gene-pair associations (n = 6). These interactions formed a compact network with discrete clusters of co-occurring alterations, suggesting that SS is characterized by a more coordinated and potentially hierarchical mutational structure. The restricted number of significant relationships indicates that a smaller subset of genetic events may cooperate to drive disease biology, consistent with a model in which key regulatory nodes, particularly those linked to transcriptional control and signaling fidelity, play a central role in shaping the SS phenotype.

By contrast, PCAECTCL demonstrated a markedly expanded interaction network, with 18 significant gene-pair associations identified. The resulting heatmap revealed a denser and more interconnected pattern of co-occurrence, with multiple genes participating in overlapping interaction clusters. This broader network architecture suggests a more heterogeneous and distributed mutational landscape, in which multiple combinations of alterations may contribute to tumor development and progression. In addition to co-occurrence, several gene pairs exhibited patterns consistent with mutual exclusivity, further highlighting the presence of alternative, potentially redundant oncogenic routes within PCAECTCL tumors.

Importantly, the magnitude and diversity of these interactions were reflected in the distribution of −log10(p-value) scores, with PCAECTCL displaying a wider range of statistically supported associations compared to SS. While SS interactions were fewer and more localized, PCAECTCL exhibited both stronger and more numerous signals, reinforcing the notion of increased combinatorial complexity.

Taken together, these findings indicate that SS and PCAECTCL differ not only in the frequency of pathway alterations but also in how somatic mutations are organized across the genome. SS appears to follow a more constrained mutational program with selective co-dependencies, whereas PCAECTCL is characterized by a more expansive and flexible network of interacting genetic events. This divergence in co-mutation architecture provides additional evidence of distinct evolutionary trajectories and may have implications for therapeutic targeting, particularly in identifying vulnerabilities within tightly coordinated versus distributed oncogenic systems.

### 3.6 Oncoplot analysis highlights subtype-specific mutational patterns and gene-level differences

To characterize the distribution of somatic alterations at the individual tumor level, we generated an oncoplot of the most frequently mutated genes across the cohort, stratified by Sézary syndrome (SS) and PCAECTCL (Fig. 3). This visualization revealed both shared and subtype-specific patterns, offering a detailed view of gene-level differences that underpin the distinct molecular architectures of these CTCL entities.

**Figure 3.**
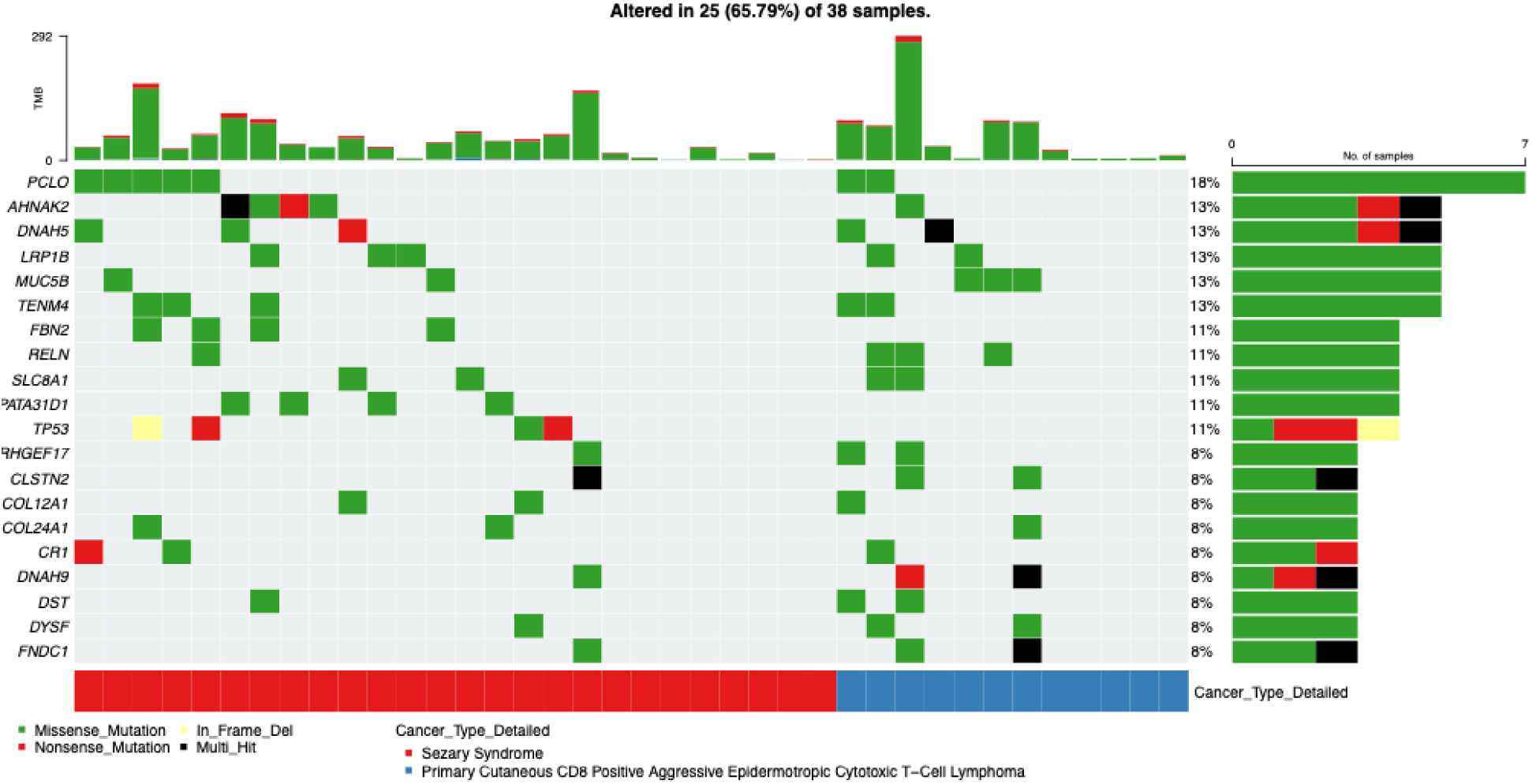
Oncoplot depicting the mutational landscape of Sézary syndrome and PCAECTCL. This oncoplot summarizes somatic mutation patterns across the 20 most frequently mutated genes identified in the cutaneous T-cell lymphoma (CTCL) cohort. Each column represents an individual tumor sample and each row corresponds to a gene, while colored tiles indicate the presence and type of high-impact somatic alterations detected in the genomic dataset. Samples are arranged by subtype, with Sézary syndrome (SS) displayed on the left and Primary Cutaneous CD8 Positive Aggressive Epidermotropic Cytotoxic T-Cell Lymphoma (PCAECTCL) on the right. The bar plot above the heatmap represents the tumor mutation burden (TMB) for each sample based on clinical annotations from the dataset. The annotation track below the heatmap indicates the detailed CTCL subtype classification for each tumor. Overall, the oncoplot highlights subtype-specific mutation patterns and heterogeneity across the cohort, illustrating differences in the distribution of recurrent gene alterations and overall mutational profiles between SS and PCAECTCL.

In the SS cohort, mutations were concentrated within a relatively defined subset of genes, with evidence of recurrent alterations across multiple samples. The most frequent mutation was observed in PCLO (19%), followed by TP53, AHNAK2, FBN2, and SPATA31D1 (each 15%). Additional low-frequency mutations were detected in genes such as FNDC1, DYSF, DST, DNAH9, CLSTN2, and ARHGEF17 (each ∼4%). These alterations appeared across several SS tumors, forming a relatively consistent recurrence pattern and suggesting a more structured mutational landscape. Functionally, many of these genes are linked to transcriptional regulation, cytoskeletal organization, and signaling processes, supporting a coordinated set of biological perturbations in SS.

In contrast, the PCAECTCL cohort exhibited a more heterogeneous and less recurrent mutational profile. The most frequently altered genes were MUC5B and RELN (each 23%), with additional mutations occurring at low frequency in genes such as CR1 and COL24A1 (each ∼8%). Unlike SS, these alterations were more sparsely distributed across tumors, with fewer genes showing repeated mutations across multiple samples. This pattern reflects a more dispersed genomic architecture, suggesting that PCAECTCL may arise through a broader range of mutational combinations rather than a consistent set of recurrent drivers.

Several genes demonstrated clear subtype-restricted patterns. Mutations in TP53, FBN2, and SPATA31D1 were observed exclusively in SS and were absent in PCAECTCL, highlighting potential subtype-specific molecular features. In contrast, no genes showed strong exclusivity for PCAECTCL within this dataset, suggesting that its mutational landscape may be defined more by diversity than by unique recurrent drivers.

The top barplot further confirmed that tumor mutation burden varied across individual samples but did not differ systematically between subtypes. This reinforces the conclusion that the primary distinction between SS and PCAECTCL lies not in mutation quantity, but in the identity, recurrence, and distribution of somatic alterations. Overall, SS is characterized by a more focused and recurrent gene-level mutational profile, whereas PCAECTCL displays greater heterogeneity and genomic dispersion, consistent with distinct underlying oncogenic programs and evolutionary trajectories.

Visualization of mutation distribution using lollipop plots indicated that alterations were broadly spread along protein-coding regions, rather than concentrated within a limited set of recurrent hotspots (Figs. S2 and S3), supporting a model of heterogeneous, pathway-driven mutagenesis in CTCL. These graphical assessments emphasize that the key differences between SS and PCAECTCL lie not in the overall number of mutations, but in the specific genes affected, their functional pathway context, and the way these alterations interact within each subtype.

### 3.7 Conversational AI-guided analyses refine subtype-specific genomic and clinical patterns

To further interrogate subtype-associated molecular features, we leveraged the conversational AI framework to perform targeted case-control analyses integrating genomic and clinical variables across Sézary syndrome (SS) and PCAECTCL cohorts. Within this workflow, AI-enabled querying facilitated rapid cohort definition, dynamic comparison of mutation prevalence, and real-time visualization of results, enabling efficient prioritization of candidate subtype-specific patterns.

We first evaluated differences in MAPK pathway alterations using an AI-guided case-control framework (Fig. S4). Comparison of SS (n = 26) and PCAECTCL (n = 13) samples demonstrated that MAPK alterations were present in both subtypes, with a modestly higher proportion observed in PCAECTCL. However, statistical testing did not reveal a significant difference (p = 0.397), and the estimated odds ratio (0.278) was associated with a wide confidence interval, reflecting the limited sample size. These findings suggest that while MAPK signaling may contribute to PCAECTCL biology, it does not represent a defining or exclusive feature distinguishing the two subtypes.

We next extended this approach to evaluate broader molecular alteration patterns (Fig. S5). Conversational AI-driven comparisons of selected genomic features showed overlapping distributions of altered and non-altered samples between SS and PCAECTCL. Consistent with pathway-level analyses, no statistically significant differences were observed for these features, reinforcing the concept that both subtypes share elements of their mutational landscape despite differences in pathway organization and gene-level recurrence.

To integrate clinical and molecular dimensions, we performed an AI-assisted evaluation of feature associations across cohorts (Fig. S6). As expected, subtype-defining clinical variables, including “Cancer Type Detailed” and “Subtype_Group,” showed highly significant differences between SS and PCAECTCL (P ≈ 3.99×10⁻⁹), validating cohort stratification. In contrast, individual gene-level variables such as MAPK1, PREX2, ARID1A, and TP53, as well as aggregated MAPK pathway status, did not demonstrate statistically significant associations with subtype classification. These results are consistent with prior observations that the distinction between SS and PCAECTCL is not driven by single-gene events but rather by coordinated differences in pathway-level architecture.

Overall, these conversational AI-guided analyses highlight the utility of this framework in rapidly generating and testing hypotheses across integrated clinical-genomic datasets. While individual gene and pathway comparisons did not consistently reach statistical significance, the overall pattern supports a model in which SS and PCAECTCL share comparable mutation frequencies but differ in how these alterations are organized and functionally integrated. By enabling iterative exploration and prioritization of biologically coherent signals, conversational AI complements traditional statistical approaches and facilitates deeper insight into subtype-specific molecular programs in CTCL.

## 4. Discussion

In this study, we performed a focused pathway-centric comparison of two aggressive but biologically distinct CTCL entities, SS and PCAECTCL, using a conversational artificial intelligence-assisted analytical framework applied to public genomic data. Several key findings emerged. First, SS and PCAECTCL did not differ in overall tumor mutational burden, indicating that the distinction between these subtypes is not explained by greater mutation quantity. Second, the two diseases diverged at the level of pathway architecture, with SS showing greater involvement of epigenetic regulators, tumor suppressor and cell-cycle programs, NFAT-associated signaling, T-cell receptor-linked pathways, and DNA damage response, whereas PCAECTCL showed relatively greater representation of MAPK pathway alterations. Third, gene-level and co-mutation analyses suggested that SS is characterized by a more focused and recurrent mutational structure, while PCAECTCL exhibits a broader and more distributed genomic interaction landscape. Together, these results support the concept that aggressive CTCL subtypes are better distinguished by the identity, organization, and functional context of their somatic alterations than by overall mutational load alone.

One of the clearest signals in our study was the relative enrichment of epigenetic regulatory alterations in SS. Recurrent SS-associated mutations involved genes such as TET2, CREBBP, CHD3, and BRD9, supporting the idea that chromatin remodeling and transcriptional deregulation are central features of this subtype. This observation is biologically plausible in light of prior CTCL studies showing recurrent disruption of epigenetic machinery and transcriptional regulators across malignant T-cell states. In SS, such alterations may be especially relevant because the disease is defined not only by skin involvement but also by systemic dissemination and circulating malignant T cells. Epigenetic dysregulation may provide a mechanistic basis for the transcriptional plasticity, immune escape, and lineage-associated reprogramming that underlie this leukemic phenotype. Rather than acting as isolated lesions, these mutations likely cooperate to stabilize aberrant transcriptional states that promote survival and persistence of malignant clones.

A second major feature of SS in our analysis was the greater involvement of tumor suppressor and cell-cycle pathways, including recurrent TP53 alterations. Although the absolute mutation frequencies were modest, the consistent direction of enrichment suggests that loss of checkpoint control may be an important component of SS biology. In the context of concurrent epigenetic dysregulation, disruption of tumor suppressor function may facilitate clonal expansion while reducing the ability of malignant cells to undergo apoptosis or senescence in response to genomic stress. This combination of transcriptional deregulation and impaired growth control provides a coherent model for the aggressive clinical behavior of SS. It also supports the broader idea that subtype-defining biology in CTCL emerges from combinations of lesions that converge on shared cellular programs rather than from a single dominant driver.

Our results also point toward aberrant T-cell activation circuitry as a more prominent feature of SS. Alterations involving PLCG1, PRKG1, CARD11, and NFAT-related pathways suggest persistent engagement of proximal T-cell receptor (TCR) and downstream transcriptional signaling. These pathways are highly relevant to malignant T-cell biology, as they regulate activation, proliferation, cytokine output, and cell fate. In SS, where malignant cells circulate and interact dynamically with multiple immune compartments, deregulation of TCR/NFAT signaling may contribute both to intrinsic tumor cell fitness and to the broader immune dysregulation that characterizes the disease clinically. The presence of low-frequency mutations in PDCD1 and other immune-related genes further raises the possibility that SS may acquire selective advantages through altered immune checkpoint signaling and impaired apoptotic regulation, even if such events are not common across all tumors.

By contrast, PCAECTCL showed a different genomic emphasis. Although epigenetic pathway alterations were also present in this subtype, the relative enrichment of MAPK-associated alterations distinguished PCAECTCL from SS. Recurrent mutations in MAPK1, PREX2, BRAF, and related signaling genes suggest that mitogenic signaling may play a more central role in at least a subset of PCAECTCL tumors. The detection of ATR and ARID1A mutations further points to contributions from DNA damage response and chromatin remodeling, but without the same recurrent and concentrated pattern observed in SS. Instead, PCAECTCL appears to be shaped by a more heterogeneous collection of lower-frequency alterations distributed across several signaling axes. This broader dispersion may reflect underlying biological complexity linked to its cytotoxic CD8⁺ phenotype, epidermotropic behavior, and highly aggressive clinical course.

The contrast between these subtypes became even more apparent in the co-mutation analyses. SS displayed a relatively constrained network with fewer significant gene-gene interactions, suggesting a more selective mutational architecture built around a narrower set of cooperating lesions. In practical terms, this may indicate that SS depends on a limited number of core biological programs, particularly those related to epigenetic regulation, T-cell signaling, and transcriptional control. PCAECTCL, in contrast, exhibited a substantially larger and denser gene-gene interaction network, with both co-occurrence and mutual exclusivity patterns. This finding implies a more flexible and distributed evolutionary structure, in which different combinations of mutations may support tumorigenesis in different cases. Such a topology is consistent with greater genomic heterogeneity and with the possibility of multiple parallel oncogenic routes converging on an aggressive phenotype.

The oncoplot findings further support this distinction in mutational architecture between SS and PCAECTCL. SS tumors exhibited recurrent alterations within a relatively constrained set of genes, most notably PCLO (19%) and TP53, AHNAK2, FBN2, and SPATA31D1 (each 15%), with additional low-frequency events distributed across a limited number of genes. In contrast, PCAECTCL tumors showed a more diffuse pattern, with mutations spread across a broader gene set and fewer recurrent events, including MUC5B and RELN (each 23%) and sporadic alterations in genes such as CR1 and COL24A1. Importantly, several genes, including TP53, FBN2, and SPATA31D1, were exclusively mutated in SS, underscoring subtype-specific molecular features. These visual patterns were consistent with the tumor mutational burden analysis, which demonstrated comparable overall mutation loads across subtypes. Taken together, these observations indicate that the key distinction between SS and PCAECTCL lies not in the number of mutations but in their recurrence, distribution, and biological context. This supports a model in which SS is driven by a more coordinated and recurrent set of genomic alterations, whereas PCAECTCL reflects a more heterogeneous and combinatorial mutational landscape, with important implications for understanding subtype-specific disease biology.

An additional finding with potential translational relevance was the identification of ERBB2 as a subtype-enriched alteration in PCAECTCL. Mutations in ERBB2 were detected in 23% of PCAECTCL tumors and were absent in the SS cohort, reaching statistical significance despite the modest sample size. This selective enrichment suggests that ERBB2 may represent a lineage- or subtype-associated molecular feature rather than a broadly shared event across CTCL. Biologically, ERBB2 encodes a receptor tyrosine kinase that plays a central role in growth factor signaling, cellular proliferation, and survival, and has been successfully targeted in several solid tumors. Its presence in PCAECTCL raises the possibility that a subset of these tumors may rely on ERBB2-mediated signaling pathways, providing a rationale for exploring targeted therapeutic strategies in this otherwise aggressive and treatment-refractory disease. At the same time, the absence of ERBB2 alterations in SS reinforces the concept of divergent oncogenic dependencies between CD4⁺ leukemic and CD8⁺ cytotoxic CTCL subtypes. While these findings require validation in larger cohorts and functional studies, they highlight how gene-level analyses can uncover clinically actionable hypotheses that may not be apparent from pathway-level summaries alone.

A distinctive aspect of this work is the incorporation of conversational AI into the genomic discovery process. In this study, AI-HOPE (26), AI-HOPE-JAK-STAT (27) and AI-HOPE-MAPK (28), functioned as analytical accelerators rather than a substitute for statistical rigor. The platform facilitated rapid hypothesis generation, cohort refinement, pathway prioritization, and iterative interpretation of results from a public dataset. This is particularly valuable in rare malignancies, where sample sizes are modest, biological heterogeneity is substantial, and exploratory analyses often require repeated reframing of questions. The utility of conversational AI in this setting lies in its ability to organize complex observations into testable biological narratives while preserving compatibility with conventional statistical workflows. As translational oncology increasingly depends on integration of multidimensional datasets, such frameworks may help bridge the gap between raw genomic output and clinically meaningful interpretation.

Several limitations should be considered when interpreting these findings. First, the study is based on a relatively small public cohort, which restricts statistical power for both gene-level and pathway-level comparisons. As a result, many of the subtype-associated patterns observed here should be interpreted as directional and hypothesis-generating rather than definitive. Second, the analysis is cross-sectional and cannot establish the temporal order of mutational acquisition or clonal evolution. Third, although we used curated pathway groupings to enhance biological interpretability, pathway assignment inevitably simplifies complex signaling relationships and may not fully capture context-dependent gene functions. Fourth, this study is based on genomic data alone and does not incorporate transcriptomic, epigenomic, proteomic, or spatial microenvironmental information, all of which are highly relevant in CTCL. Finally, functional validation was beyond the scope of this work, and the mechanistic relevance of the identified alterations will require experimental confirmation.

Despite these limitations, the study provides several important conceptual advances. By restricting the comparison to SS and PCAECTCL, we reduced subtype heterogeneity and enabled a cleaner evaluation of lineage- and phenotype-associated molecular differences than would be possible in broader pooled “non-SS” groups. This focused design revealed that SS and PCAECTCL are not simply variants within a shared mutational continuum; rather, they appear to represent distinct pathway-organized disease states. SS is dominated by recurrent alterations in epigenetic, transcriptional, and immune-regulatory circuitry, whereas PCAECTCL shows a more heterogeneous genomic architecture with relative MAPK pathway prominence and a more complex interaction network. These distinctions may help explain differences in clinical presentation and could inform future efforts to develop subtype-adapted therapeutic strategies.

Future work should extend these observations in several directions. Larger multi-institutional cohorts will be necessary to validate subtype-specific genes and pathway enrichments, especially for rare entities such as PCAECTCL. Integration with transcriptomic and single-cell datasets may clarify whether the pathway-level alterations identified here correspond to distinct malignant cell states or microenvironmental programs. Functional studies will be needed to determine whether SS-associated alterations in epigenetic and TCR/NFAT-related genes create tractable therapeutic dependencies, and whether PCAECTCL tumors with MAPK-oriented alterations exhibit sensitivity to pathway-directed inhibition. More broadly, applying conversational AI frameworks to multimodal CTCL datasets may accelerate identification of clinically actionable molecular subclasses that are not evident from histopathology alone.

In conclusion, our findings indicate that Sézary syndrome and PCAECTCL are best distinguished not by greater mutational burden, but by different pathway-level architectures, recurrent gene-level signatures, and contrasting co-mutation topologies. SS is marked by a more focused mutational program centered on epigenetic dysregulation, transcriptional control, and immune-related signaling, whereas PCAECTCL exhibits a broader and more heterogeneous network of oncogenic alterations with relative MAPK pathway prominence. These results advance the molecular understanding of aggressive CTCL and provide a rationale for subtype-specific biological investigation. They also demonstrate the value of conversational AI as a scalable companion tool for translational cancer genomics, particularly in rare and heterogeneous malignancies where extracting coherent biological signals from limited datasets remains a major challenge.

## Data Availability

All data used in the present study is publicly available at https://www.cbioportal.org/ and https://genie.cbioportal.org. The datasets used in our study were aggregated/summary data, and no individual-level data were used. Additional data can be provided upon reasonable request to the authors.

---

**Figure S1.**
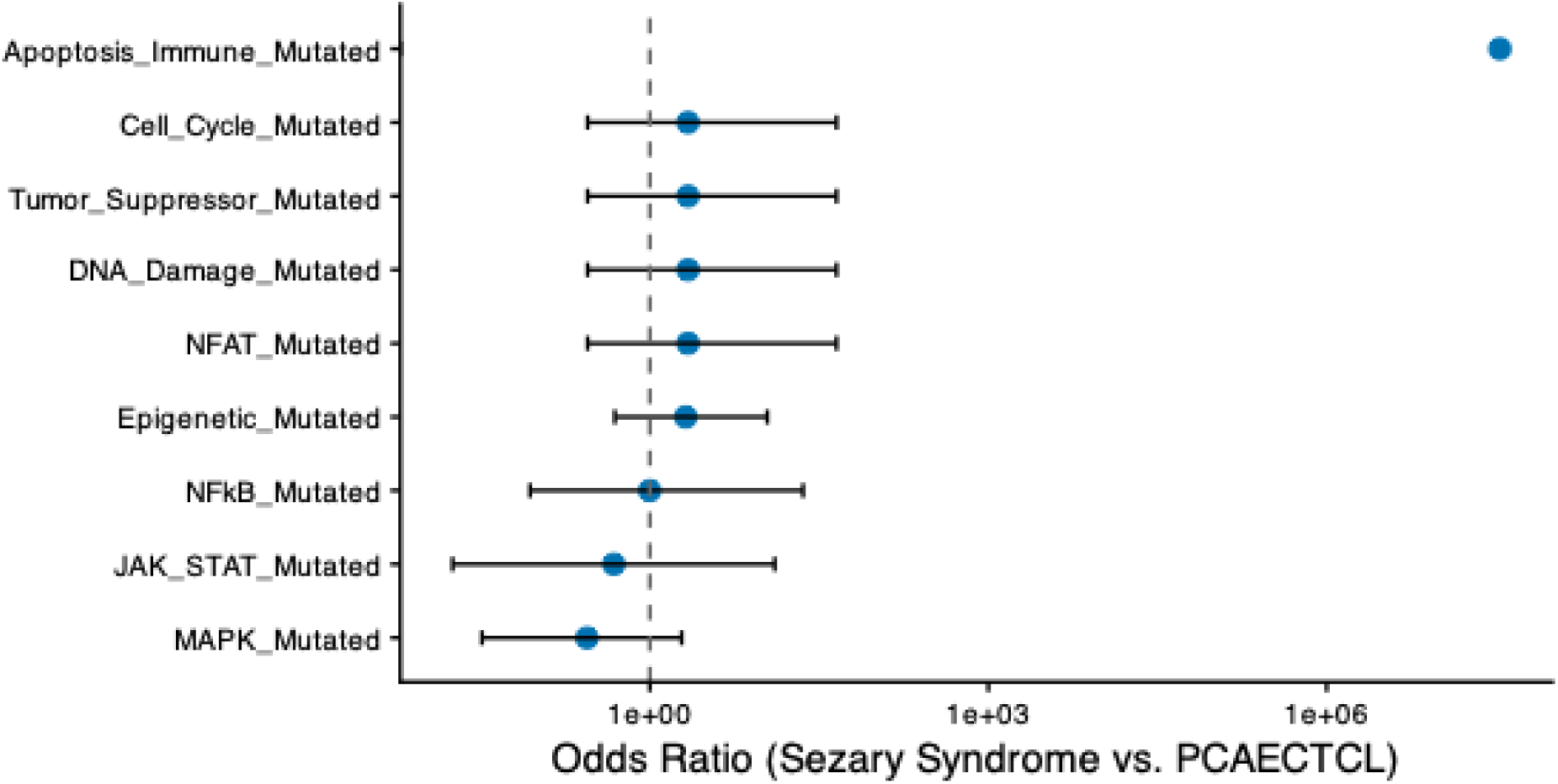
Comparative odds ratios of pathway-level alterations between Sézary syndrome and PCAECTCL. This forest plot displays the odds ratios and associated confidence intervals for pathway-level mutation frequencies comparing Sézary syndrome (SS) to primary cutaneous CD8⁺ aggressive epidermotropic cytotoxic T-cell lymphoma (PCAECTCL). Values greater than one indicate relative enrichment of pathway alterations in SS, whereas values less than one reflect higher frequencies in PCAECTCL. Pathways related to epigenetic regulation, tumor suppressor function, cell-cycle control, NFAT signaling, and DNA damage response demonstrate a consistent shift toward higher representation in SS, suggesting a greater contribution of transcriptional and regulatory disruption in this subtype. In contrast, MAPK and JAK/STAT signaling pathways trend below unity, indicating relatively increased involvement in PCAECTCL. NF-κB pathway alterations cluster near the null value, consistent with comparable frequencies across both cohorts. Notably, apoptosis and immune regulatory pathways show the largest deviation toward SS, driven by their presence in SS tumors and absence in PCAECTCL. The distribution of odds ratios highlights distinct pathway-level biases between these aggressive CTCL subtypes, reinforcing differences in their underlying molecular programs.

**Figure S2.**
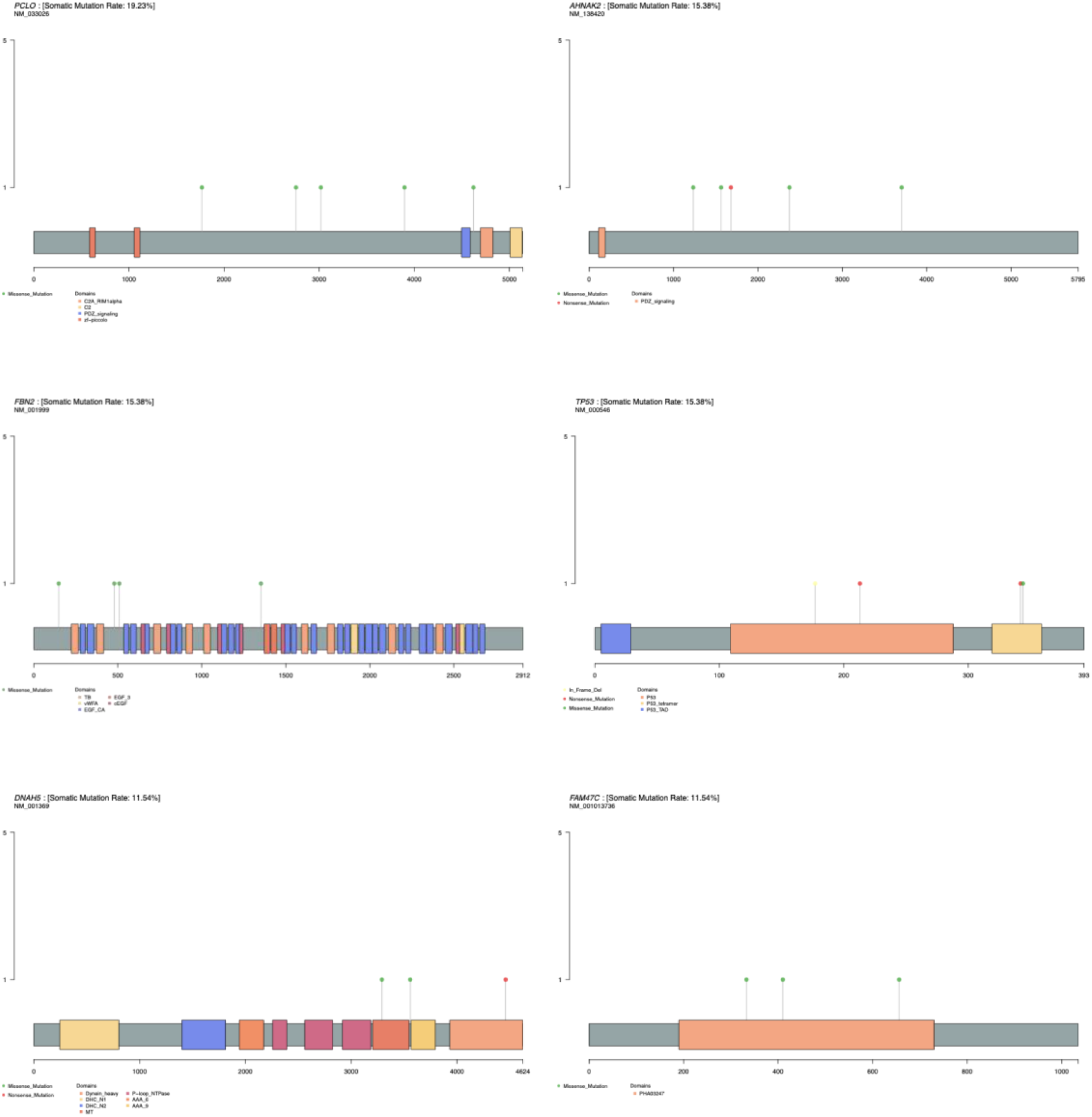
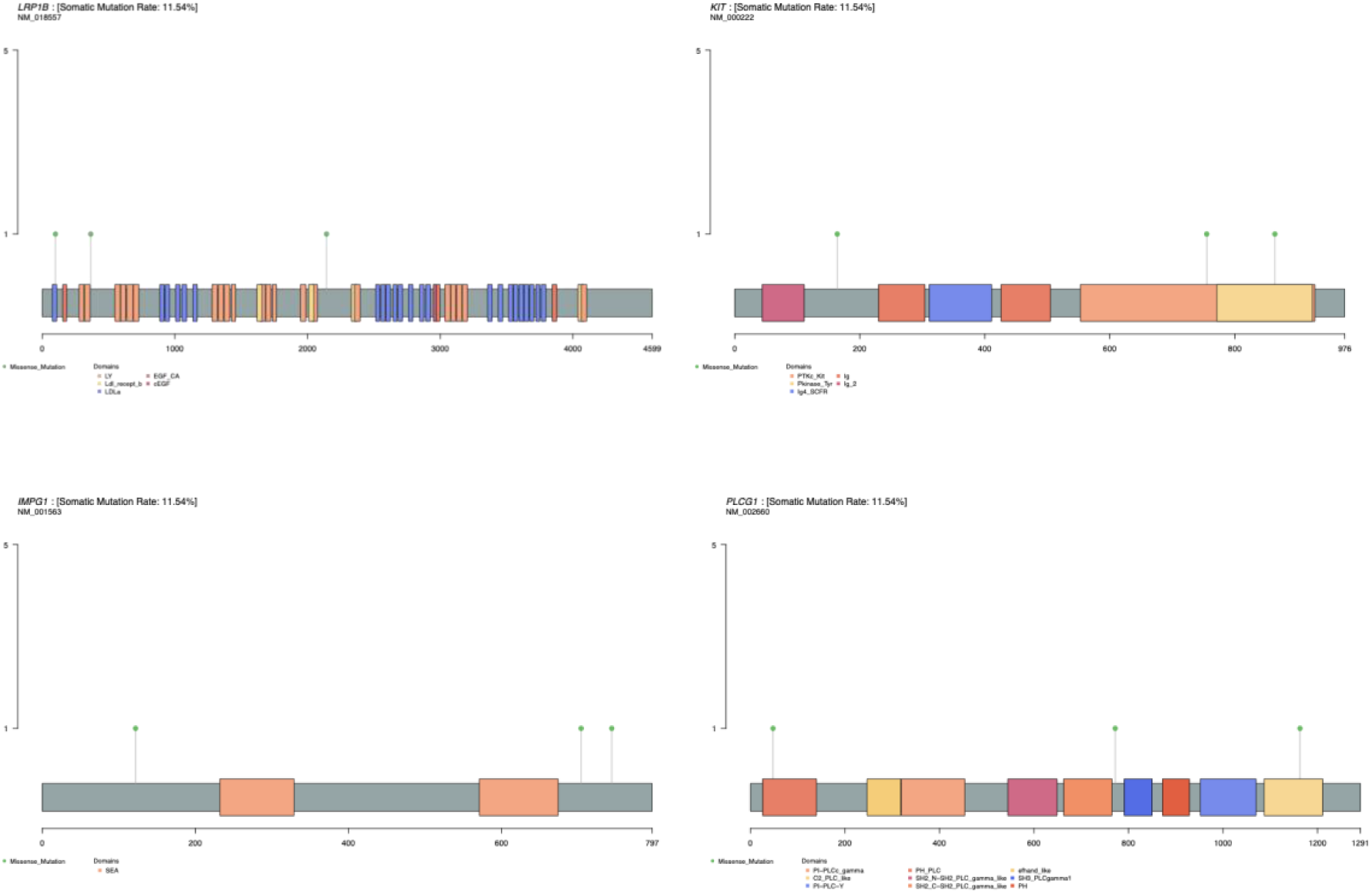
Lollipop plots illustrating the distribution of somatic mutations in the top 10 recurrently mutated genes in the Sézary syndrome cohort. This figure presents lollipop plots depicting the positional distribution of somatic mutations across the protein sequences of the ten most frequently mutated genes identified in the Sézary syndrome (SS) cohort. Each panel corresponds to a single gene, and each lollipop marker represents an individual mutation event. The horizontal axis denotes the amino acid position along the protein sequence, while the vertical lollipop height reflects the number of tumor samples harboring mutations at that site. Colored markers indicate the type of somatic alteration, and annotated protein domains are shown along the protein structure to contextualize mutation locations. Together, these plots highlight mutation clustering and potential hotspot regions across the most frequently altered genes in SS, providing insight into candidate functional domains that may contribute to disease pathogenesis.

**Figure S3.**
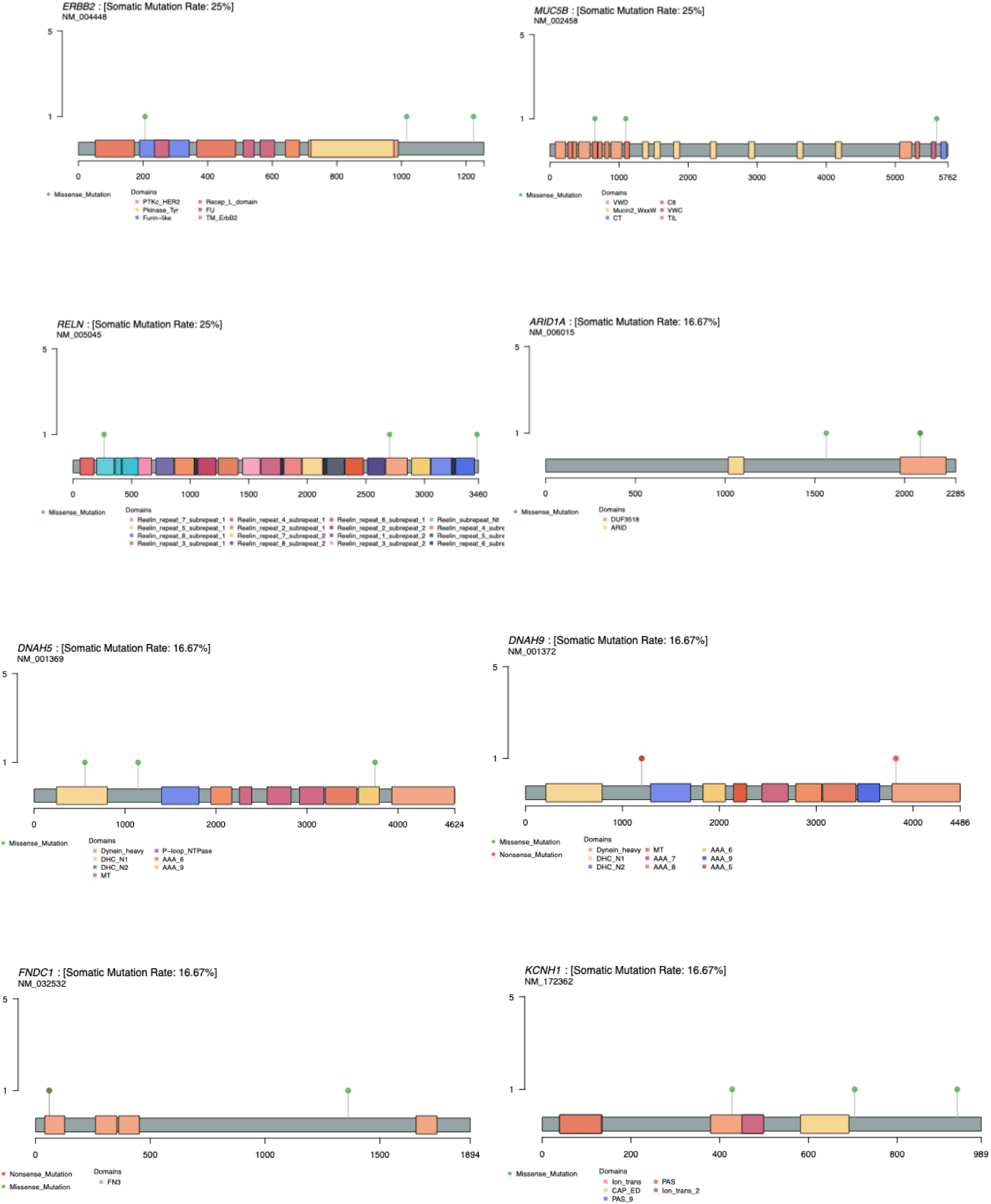
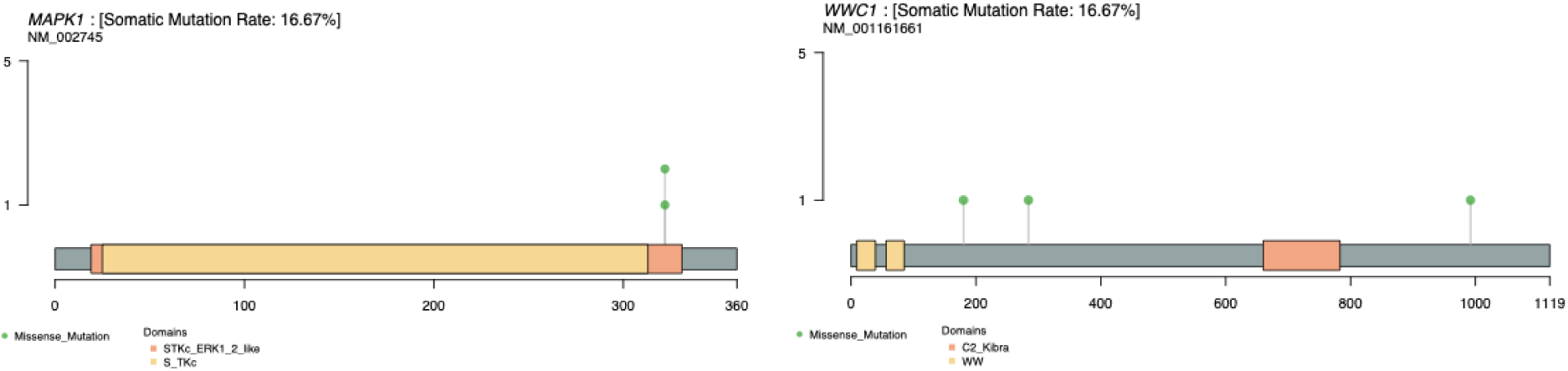
Lollipop plots illustrating the distribution of somatic mutations in the top 10 recurrently mutated genes in the Primary Cutaneous CD8⁺ Aggressive Epidermotropic Cytotoxic T-Cell Lymphoma (PCAECTC) cohort. This figure presents lollipop plots depicting the positional distribution of somatic mutations across the protein sequences of the ten most frequently mutated genes identified in the PCAECTC. Each panel corresponds to an individual gene, with lollipop markers representing mutation events observed across tumor samples. The horizontal axis indicates the amino acid position along the protein sequence, while the vertical height of each lollipop reflects the number of samples harboring mutations at that specific site. Colored markers denote different mutation types, and annotated protein domains provide structural context for the observed alterations. These visualizations highlight mutation distribution patterns and potential hotspot regions across recurrently altered genes in PCAECTC, offering insight into candidate functional regions that may contribute to disease biology.

**Figure S4.**
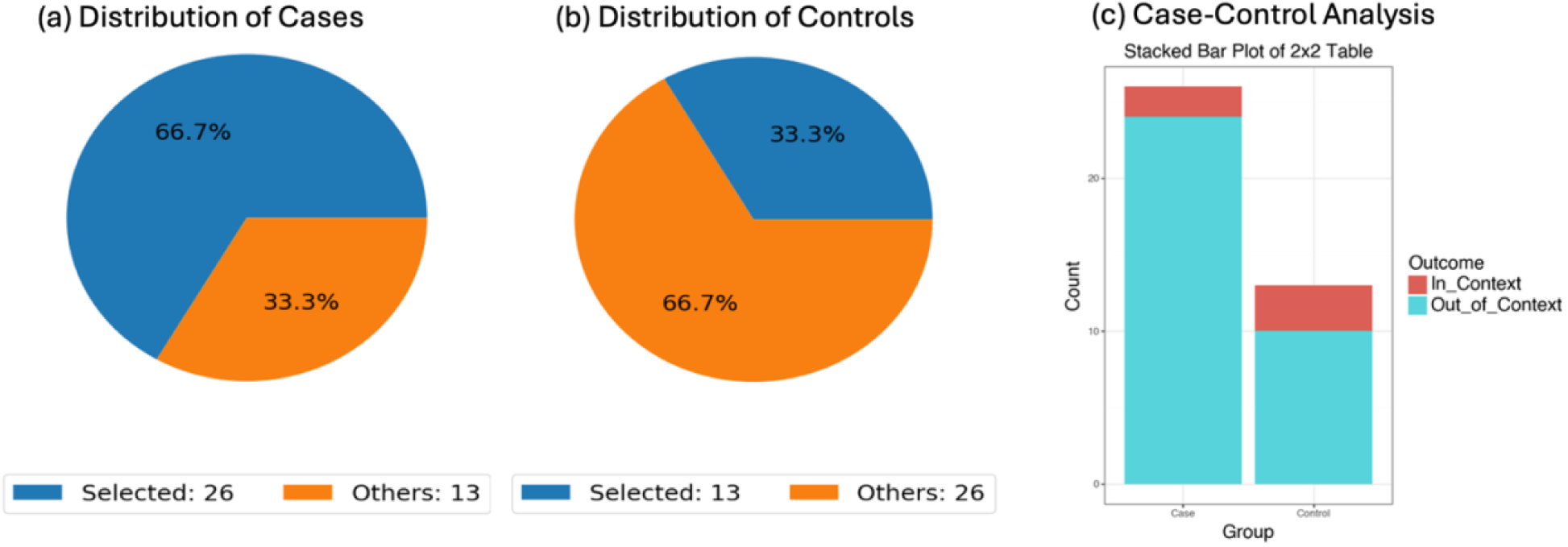
Conversational AI-guided case-control evaluation of MAPK pathway alterations in Sézary syndrome and PCAECTCL. This figure presents a conversational AI-assisted analytical workflow designed to assess differences in the prevalence of MAPK pathway alterations between Sézary syndrome (SS) and primary cutaneous CD8⁺ aggressive epidermotropic cytotoxic T-cell lymphoma (PCAECTCL). The case cohort consisted of SS samples (n = 26), while the control cohort included PCAECTCL samples (n = 13), defined through structured query logic within the AI-driven platform. Panels (a) and (b) display the proportional representation of selected samples relative to the full dataset for each cohort, illustrating the relative cohort sizes and composition. Panel (c) summarizes the distribution of MAPK-mutated (“in-context”) and non-mutated (“out-of-context”) samples using a stacked bar plot derived from a 2×2 contingency framework. MAPK alterations were identified in a subset of tumors in both groups, with a slightly higher proportion observed in PCAECTCL compared to SS. Statistical evaluation yielded a non-significant association (p = 0.397), with an odds ratio of 0.278 and a wide confidence interval, reflecting limited statistical power. Overall, this analysis demonstrates how conversational AI frameworks can be leveraged to rapidly construct and visualize subtype-specific comparisons of pathway alterations, supporting exploratory interrogation of genomic differences across CTCL subtypes.

**Figure S5.**
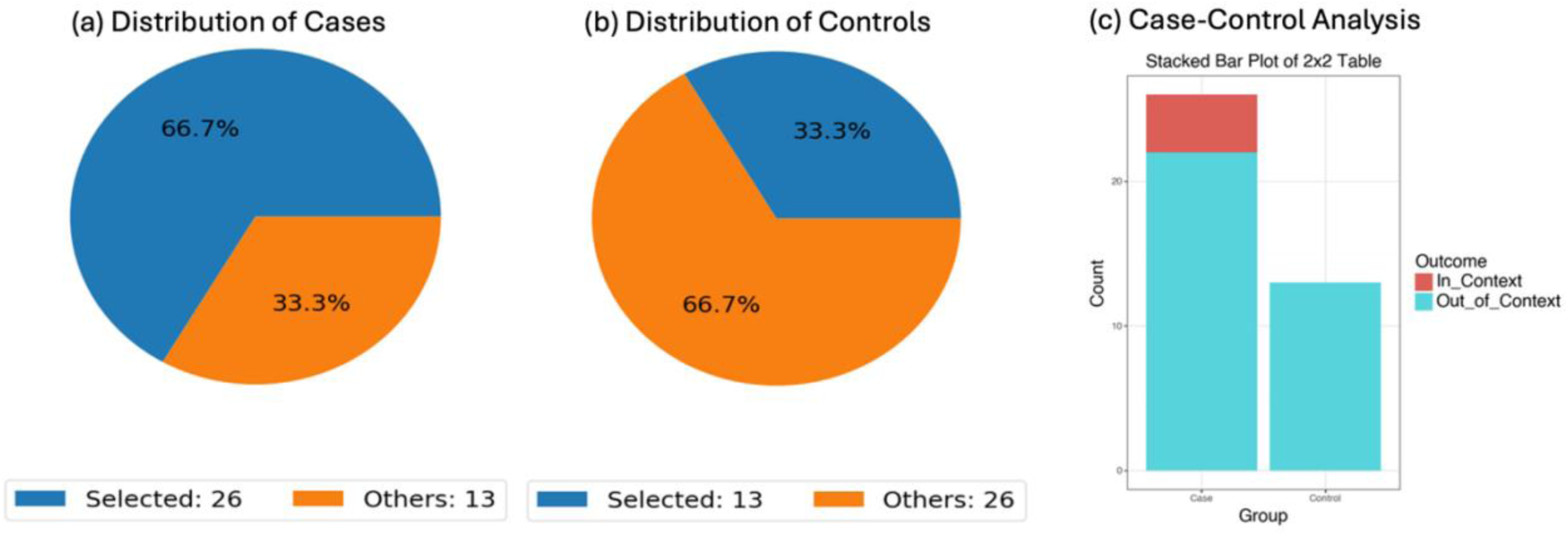
Conversational AI-enabled case-control assessment of pathway alteration prevalence between Sézary syndrome and PCAECTCL. This figure illustrates a conversational AI-driven case-control analysis evaluating differences in the prevalence of selected molecular alterations between Sézary syndrome (SS) and primary cutaneous CD8⁺ aggressive epidermotropic cytotoxic T-cell lymphoma (PCAECTCL). Cohorts were defined within the AI framework using structured query criteria, with SS samples designated as the case group (n = 26) and PCAECTCL samples as the control group (n = 13). Panels (a) and (b) depict the proportional representation of selected samples relative to the full dataset for each cohort, highlighting the relative sizes and composition of the groups included in the analysis. Panel (c) presents a stacked bar plot summarizing the number of samples with (“in-context”) and without (“out-of-context”) the specified molecular alteration using a 2×2 contingency structure. The distribution of altered versus non-altered samples appears comparable between cohorts, and statistical testing did not reveal a significant difference in prevalence, consistent with overlapping mutation frequencies. This visualization demonstrates how conversational AI frameworks can rapidly construct and interpret case-control comparisons, enabling efficient exploration of subtype-specific genomic patterns in CTCL.

**Figure S6.**
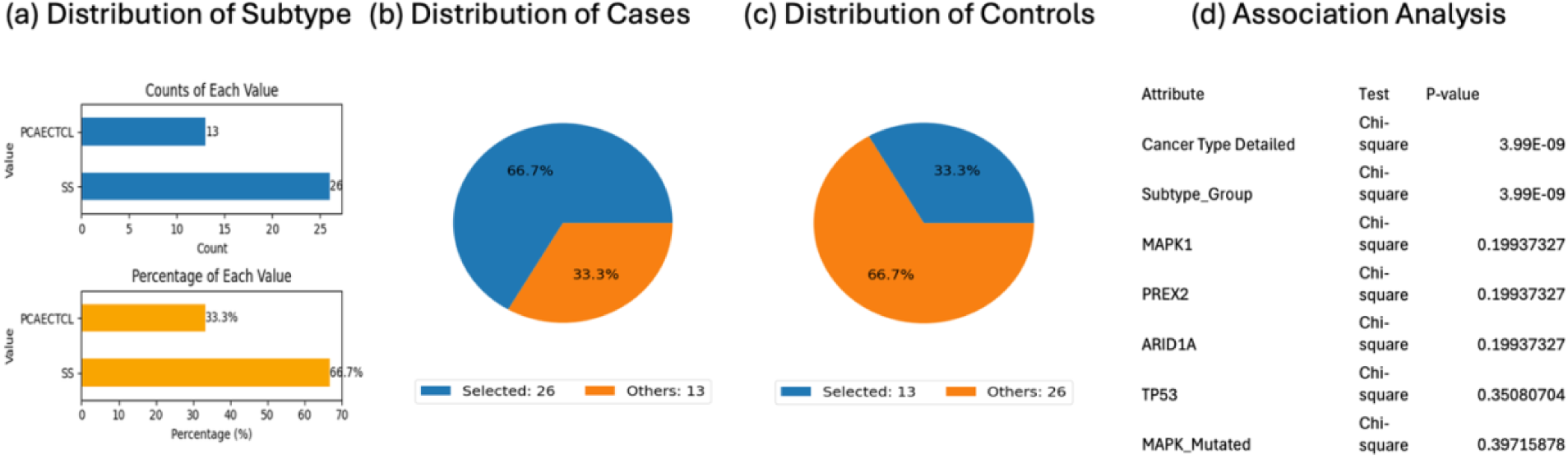
Conversational AI-assisted analysis of clinical and molecular feature associations between Sézary syndrome and PCAECTCL. This figure summarizes a conversational AI-driven case-control framework used to explore associations between clinical attributes and selected genomic features in Sézary syndrome (SS) and primary cutaneous CD8⁺ aggressive epidermotropic cytotoxic T-cell lymphoma (PCAECTCL). The case cohort consisted of SS samples (n = 26), while the control cohort included PCAECTCL samples (n = 13), defined using structured query logic within the AI platform. Panel (a) displays the distribution of subtype classification across the dataset, illustrating both absolute counts and relative proportions for each group. Panels (b and c) presents pie charts depicting the proportion of selected versus non-selected samples within the case and control cohorts, reflecting their contribution to the overall dataset. Panel (d) summarizes statistical associations between clinical variables and molecular features using chi-square testing. As expected, subtype-defining variables such as “Cancer Type Detailed” and “Subtype_Group” show highly significant differences between cohorts (P ≈ 3.99×10⁻⁹). In contrast, individual gene-level features, including MAPK1, PREX2, ARID1A, TP53, and aggregated MAPK pathway status, do not demonstrate statistically significant differences, consistent with the broader observation of comparable mutation burdens but divergent pathway organization. Overall, this figure highlights how conversational AI can integrate clinical and genomic variables to systematically evaluate subtype-associated patterns in CTCL.

**Table S1.** Gene-level comparison of borderline-significant somatic mutation frequencies between Sézary syndrome and PCAECTCL cohorts.

| Gene | Sézary Syndrome Mutated n (%) | Sézary Syndrome Wild-Type n (%) | PCAECTCL Mutated n (%) | PCAECTCL Wild-Type n (%) | p-value |
| --- | --- | --- | --- | --- | --- |
| ERBB2 | 0 (0%) | 26 (100%) | 3 (23%) | 10 (77%) | **0.03129** |

## Notes

### Competing Interest Statement

The authors have declared no competing interest.

### Funding Statement

This study was funded by the National Cancer Institute, NCI, award number U2CCA252971; the City of Hope Cancer Control and Population Sciences program by the National Institutes of Health, NIH, National Cancer Institute, NCI, award number P30CA033572; and the Drug Development and Capacity Building: A UCR/CoH-CCC Partnership project by the National Institutes of Health, NIH, National Cancer Institute, NCI, award number U54 CA285116.

### Author Declarations

The source data used in this study were publicly available before the initiation of the study and can be accessed through cBioPortal for Cancer Genomics at https://www.cbioportal.org/. Additional data may be provided upon reasonable request to the authors.

